# Long term positivity of SARS-CoV-2 total immunoglobulins in convalescent plasma and blood donors

**DOI:** 10.1101/2021.06.24.21259079

**Authors:** M Carmen Martín, Ana Jimenez, Nuria Ortega, Alba Parrado, Isabel Page, M Isabel González, Lydia Blanco-Peris

## Abstract

**Background:** One of the most questioned issues about SARS-CoV2 immunity is how long does it last. Whether lasting differences exist between infection and vaccination boosted immunity is yet to be known. The answer to this question will determine key issues such as the reliability of individual and herd immunity or the need of sanitary restrictions or periodical revaccination. The aim of this study was to determine how long total anti SARS-CoV2 antibodies due to past infection persist in peripheral blood and whether sex, age or haematological features can influence their lasting.

**Material and Methods:** A total of 2432 donations SARS-CoV-2 from 662 repeat donors from April 2020 to February 2021 were analysed. Donors were 69.7% males and their average age was 46. An automated chemilumiscence immunoassay for total antibodies recognizing N protein of SARS-CoV-2 in human serum and plasma was performed.

**Results and discussion:** In 97.6% donors with follow-up, anti SARS-CoV-2 protein N total antibodies remained positive up to 46 weeks after first positive determination. Blood group was not related to antibody waning. Lower lymphocyte counts and higher neutrophils and as well higher seric IgA would help predict future negativization of antibodies. The vast majority of donors keep their total immunoglobulins anti SARS-CoV-2 positive for longer than 10 months. Ageing might have a protective effect against antibody waning but, given the small number of cases that become negative, more studies, or larger cohorts would be needed to confirm these facts.

## Introduction

One of the most questioned issues about SARS-CoV2 immunity is how long does it last. Although evidence exists of SARS-COV-2 infection inducing long-lived bone marrow specific plasma cells even in mild cases [1], it’s not yet established whether infection or vaccination boosted immunity will last as long. The answers to this question would determine key issues such as the reliability of individual and herd immunity or the need of sanitary restrictions or periodical revaccination. Most SARS-CoV-2 infected individuals develop an effective immune response, leading to viral eradication and the production of specific T cell responses and antibodies against SARS-CoV-2 that are usually detectable 10–21 days after infection [2,3].

Antibody assays are not equivalent: they either detect antibodies against different viral proteins (S1, S1/S2, RBD or NC) or different immunoglobulin classes: IgG, IgM, IgA or their combinations. Many factors can influence test performance, including cross-reactivity with other coronaviruses, that can occur in up to 28% individuals [2] or platform (laboratory-based vs point-of-care, lateral flow). Chemiluminiscence assays have suitable performances regarding both sensibility and sensitivity and correlate with titres of neutralizing antibodies [4].

Antibody responses against viral S and N proteins are equally sensitive in the acute phase of infection, but antibody responses against N seem to wane in the post infection phase whereas those against the S protein would persist over time with a slow decay [1, 5]

Even if antibody levels waned, long-lived mononuclear blood cells (MBCs) and long-lived bone marrow plasma cells would remain to mediate rapid antibody production triggered by a new contact with the virus. Some studies suggest that SARS-CoV-2 infection strengthens pre-existing broad coronavirus protection through S2-reactive antibody and MBCs formation [6]

Antibody evolution surveys are most useful to monitor the pandemic, both before and after vaccination strategies [7]. There are several features that should be taken into account, including time lapse between infection and antibody development or antibody waning. Blood donors constitute a representative subset of population aged 18-65 and are reasonably free of biases that could over represent symptomatic or exposed individuals.

Heterogeneity of susceptibility and transmission is hard to evaluate but does exist [8]. A portion of the population may have pre-existing immunity via cross-reactivity or particular host factors such as mucosal immunity or trained innate immunity protection (as it has been reported to be conferred by Diphtheria-pertussis-tetanus (DPT) or Bacillus Calmette–Guérin (BCG) vaccination [9]. There is as well a proportion of post infection seronegative individuals that develop immunity by T cell mediated responses but without exhibiting an antibody response [10].

To date it is not possible to ascertain whether antibodies can last longer than the time COVID-19 has been among us. There are recently described early cases in France [10], arising the question of whether the pathogen could have been circulating even before the recording of the first cases in Spain by January 31^st^, 2020. Our fist positive donations are from 2019, possibly due to cross-reaction to previous coronaviruses [11] but we decided to start our series on January 13^th^, the donation date of our first positive after WHO issued a comprehensive package of technical guidance online with advice to all countries on how to detect, test and manage potential cases.

Our starting hypothesis was that the vast majority of either recovered or asymptomatic SARS-CoV-2 cases would develop and keep antibodies against the novel coronavirus [1, 2, 9]. They will subsequently contribute to herd immunity. Anyone holding antibodies due to prior infection, should be kept from receiving the second dose of vaccines requiring booster, so far their cellular response could even result impaired [12].

The main aim of this study was to determine which percentage of the seropositive population due to natural infection keeps total antibodies recognizing SARS-CoV2 and how long can they last. A secondary goal was to establish whether sex, age, blood group or haematological features might influence the fact of keeping or loosing circulating antibodies. This knowledge would help optimize immunization strategies, and would ease decisions about the need of periodical revaccination either overall or for determined population groups by reliably knowing the lasting of specific humoral immunity.

## METHODS

The aim of this study was to determine how long anti SARS-CoV2 antibodies persisted in peripheral blood, and whether haematological o demographical parameters would be related to it. A retrospective testing and information compilation was performed

Plasma-EDTA and serum frozen samples from blood, plasma (normal and convalescent) and platelet donations were tested. A total of 2432 aleatorized samples of 662 donors over 18 years old were included. Donation is allowed after one month after the end of COVID-19 symptoms in our country. The Biobank of the Centro de Hemoterapia de Castilla y León is included in the National Registry of Biobanks (RD17 / 16/2011) with the number B.0000264 and holds an ISO 9001: 2015 certification endorsing our granting of safety and traceability of any human biological sample we distribute, always behaving Spanish and European rules on human samples and data protection management.

All haematological and demographic data were extracted from electronic database eDelphyn (Hemasoft). Variables analyzed included age, sex, blood group, and laboratory data: leukocyte (WBC), neutrophil, lymphocyte, platelet, monocyte, eosinophil and basophil counts (cells*10^3^/µL) and their percentages, serum immunoglobulins IgG, IgA and IgM (mg/dL), haemoglobin (Hb) and haematocrit (HCT).

An automated chemiluminiscence double-antigen sandwich immunoassay for the in vitro semi quantitative detection of total antibodies to SARS-CoV-2 in human serum and plasma was performed. The target antigen of this immunoassay is a recombinant nucleocapsid (N) protein. Elecsys® Anti-SARS-CoV-2 (Roche Diagnostics, Basel, Switzerland) detects antibodies correlating with virus-neutralizing ones and is therefore useful to help characterize the immune reaction to SARS-CoV-2 [9, 10]. Immunoassay was validated by our serology lab by testing of 6 pairs of samples (plasma EDTA and serum) from diagnosed PCR-positive, symptomatic cases infected by mid-April, that were previously reported positive by the Spanish National Microbiology Centre, and checked to be as well positive for IgG (Chemiluminiscence, N protein, Abbott Alinity S, Chicago USA) and IgA (ELISA, S protein, Euroimmunn, Lübeck, Germany) antiSARS-CoV-2. Another set of ten prepandemic samples, therefore supposed to be negative, were equally analysed. 405 donations were analysed both in serum and plasma to verify interchangeability. A 100% concordance was yielded by all these validation assays. The cut-off was that recommended by manufacturer (OD>1 to report reactivity). Researchers performing anti SARS-CoV-2 analyses were blind to the condition of COVID-19 convalescence and to any other characteristic of the donors or to the donation dates.

Demographic and clinical characteristics of patients are expressed as their mean and standard deviation (SD) for continuous variables and frequency distributions are reported for categorical variables. Age was analysed both as continuous and categorical variable; in the latter case was recoded into 4 groups: under 30, ≥30 and <45, ≥45 and <60, and, last, ≥60 and >75 years old.

Donors whose second OD were under 90% of the first of that of their first positive were recoded into decay group. Donors over 110% were recoded as rise and those who were +/-10% of the first one were in the stay group.

Kolmogorov-Smirnov test was performed on each continuous variable to contrast normality. None of the variables followed normal distributions, therefore non-parametric Mann Whithney U test was performed to compare laboratory values. To contrast the Ho of independence within categorical variables, Pearson’s Chi-square and Fisher’s exact test were carried out. Any test was considered to be significant at a 95% confidence level.

This study was conducted according with national regulations, institutional policies and in the tenets of the Helsinki Declaration, it was approved by the Valladolid Health Area Drug Research Ethics Committee, in a meeting held on June 11th, 2020 with the reference number “BIO-2020-93”. Included donors consented to participate in Biobank research activities. The privacy rights of human subjects were always observed.

## Results

A total of 2432 donations (either whole blood, plasmapheresis or platelet apheresis) were tested for total anti SARS-CoV2 antibodies recognizing N protein. Donors made up 662; 69.7% males, aged 18-75, average 46+/-12, 46.3% A, 43.1% O, 6.5% B, 4.1 % AB. Donations were selected at random among those collected from 13^th^ January 2020 (date of the first 2020 positive found) to 11^th^ February 2021.

250 donors were positive at least once (provided a second donation after the positive one was analyzed) and therefore their 340 donations selected for further analysis. 62 donors had more than 2 positive donations. Average time gap between samples was 12.31 weeks (1-46, SD=10.01).

We considered the first positive sample of the donor within the assessed period as first donation of this study. Out of the 250 initially positive donors, only 6 (2.4%) became negative at any moment of their follow-up (Figure 1).

**Figure 1.**
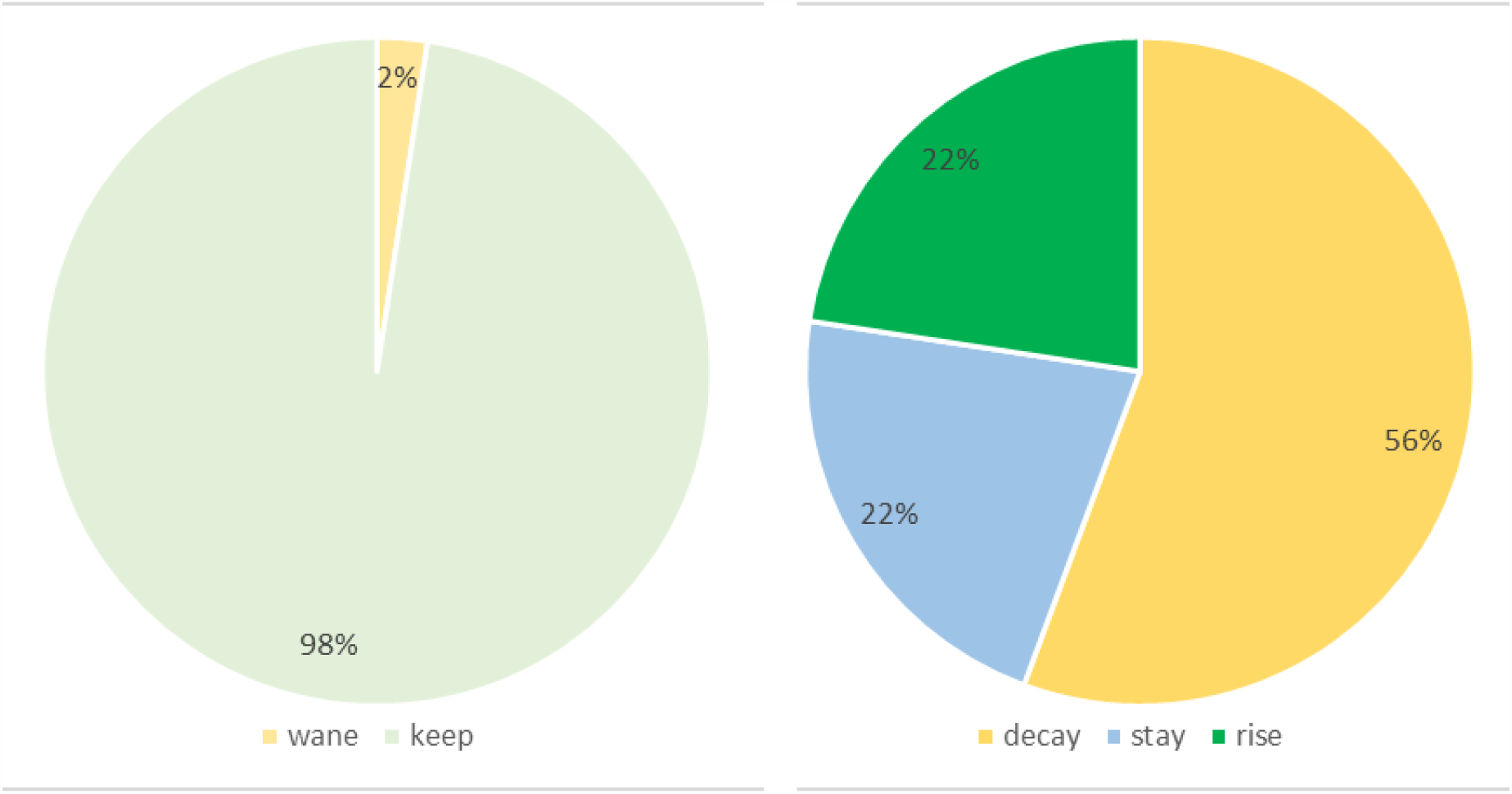
Outcomes of anti SARS-CoV-2 total antibodies (Wane: first donation positive, second, negative; decay: OD of second donation under 90% of the first one; stay: OD of second donation +/-10% of the first one; rise: OD of second donation over 110% of the first one)

**Figure 2.**
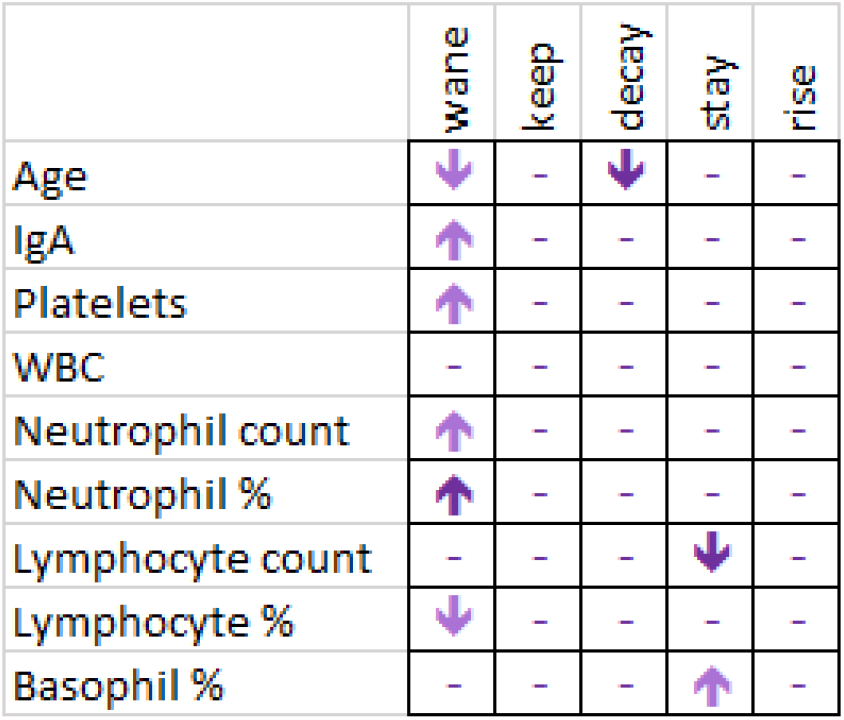
Significant changes in hematological parameters related to changes in anti SARS-CoV-2 total antibodies (wane: first donation positive, second, negative; decay: OD of second donation under 90% of the first one; stay: OD of second donation +/-10% of the first one)

Only 6 out of 250 donors lost their antibodies, all of them after just one positive donation. They were two males in their fifties and four women, two in their fifties again and two younger ones (Table1). No other wane events were found. All waning cases were weak positives (OD <2.5) in their first positive donation.

**Table 1.** Characteristics of donors loosing anti SARS-CoV-2 antibodies, optical density (OD) and interval between positive and negative donations (weeks) with total immunoglobulin to SARS-CoV2 becoming negative (OD1: optical density of first positive donation, OD2 optical density of consecutive donation to the first positive one)

| Sex | n | Age group | Blood group | DO1<br>(mean(SD)) | DO2<br>(mean(SD)) | Weeks<br>(mean(SD)) |
| --- | --- | --- | --- | --- | --- | --- |
| Male | 2 | ≥45 and <60 | A, O | 1.51 (0.59) | 0.55 (0.05) | 12 (1.41) |
| Female | 1 | under 30 | A | 1.05 (-) | 0.88 (-) | 26 (-) |
|  | 1 | ≥30 and <45 | A | 1.08 (-) | 0.56 (-) | 14 (-) |
|  | 2 | ≥45 and <60 | O | 1.06 (0.02) | 0.72 (0.23) | 20 (8.49) |

97.6% donors kept positive their total antibodies against SARS-CoV-2. The longest period lapse between first and last donation was 46 weeks. No relationship with age, blood group or sex was found. Neither was found as comparing raw OD values.

No relationship of waning or OD changes due to blood group (Table 2) was found. Rising OD was less frequent within the under 30 group as a trend (OR=0.4; CI (0.13-1.21); p=0.149). Waning was more frequent in women as a trend (OR=3.56; CI (0.64-19.8)).

**Table 2.** Donor changes in seropositivity and OD for anti SARS-CoV-2 total immunoglobulins by sex, blood group and age group.

|  |  | Fact |  |  |  | Trend |  |  |  |  |  |
| --- | --- | --- | --- | --- | --- | --- | --- | --- | --- | --- | --- |
|  |  | wane |  | keep |  | decay |  | rise |  | stay |  |
|  |  | n | % | n | % | n | % | n | % | n | % |
| Gender | Female | 4 | 4,4% | 87 | 95,6% | 50 | 54,9% | 22 | 24,2% | 19 | 20,9% |
|  | Male | 2 | 1,3% | 157 | 98,7% | 90 | 56,6% | 32 | 20,1% | 37 | 23,3% |
| Group | A | 3 | 2,4% | 124 | 97,6% | 75 | 59,1% | 22 | 17,3% | 30 | 23,6% |
|  | AB | 0 | 0,0% | 16 | 100,0% | 8 | 50,0% | 5 | 31,3% | 3 | 18,8% |
|  | B | 0 | 0,0% | 17 | 100,0% | 9 | 52,9% | 5 | 29,4% | 3 | 17,6% |
|  | O | 3 | 3,3% | 87 | 96,7% | 48 | 53,3% | 22 | 24,4% | 20 | 22,2% |
| Age | under 30 | 1 | 2,8% | 35 | 97,2% | 25 | 69,4% | 4 | 11,1% | 7 | 19,4% |
|  | 30 to 45 | 1 | 1,2% | 80 | 98,8% | 49 | 60,5% | 17 | 21,0% | 15 | 18,5% |
|  | 45 to 60 | 4 | 3,3% | 117 | 96,7% | 60 | 49,6% | 32 | 26,4% | 29 | 24,0% |
|  | 60 to 75 | 0 | 0,0% | 12 | 100,0% | 6 | 50,0% | 1 | 8,3% | 5 | 41,7% |

Haemogram parameters (leukocytes, neutrophils, lymphocytes, monocytes, eosinophils, basophils, haematocrit and haemoglobin) from each donor were tested for its relationship with antibody waning (Table 2). Lymphocyte percent was significantly lower (25.4% vs. 31.7%; p=0.021) conversely to neutrophil percent (65.1 vs 59.1; p=0.024) in those donors whose antibodies would wane. IgA was found to be higher as a trend (324 vs. 227 mg/dL; p=0.153), as well as platelets (279.7 vs. 247.1; p=0.166) and neutrophil count (5.2 vs 4.2; p=0.129) (Table 3).

**Table 3.**
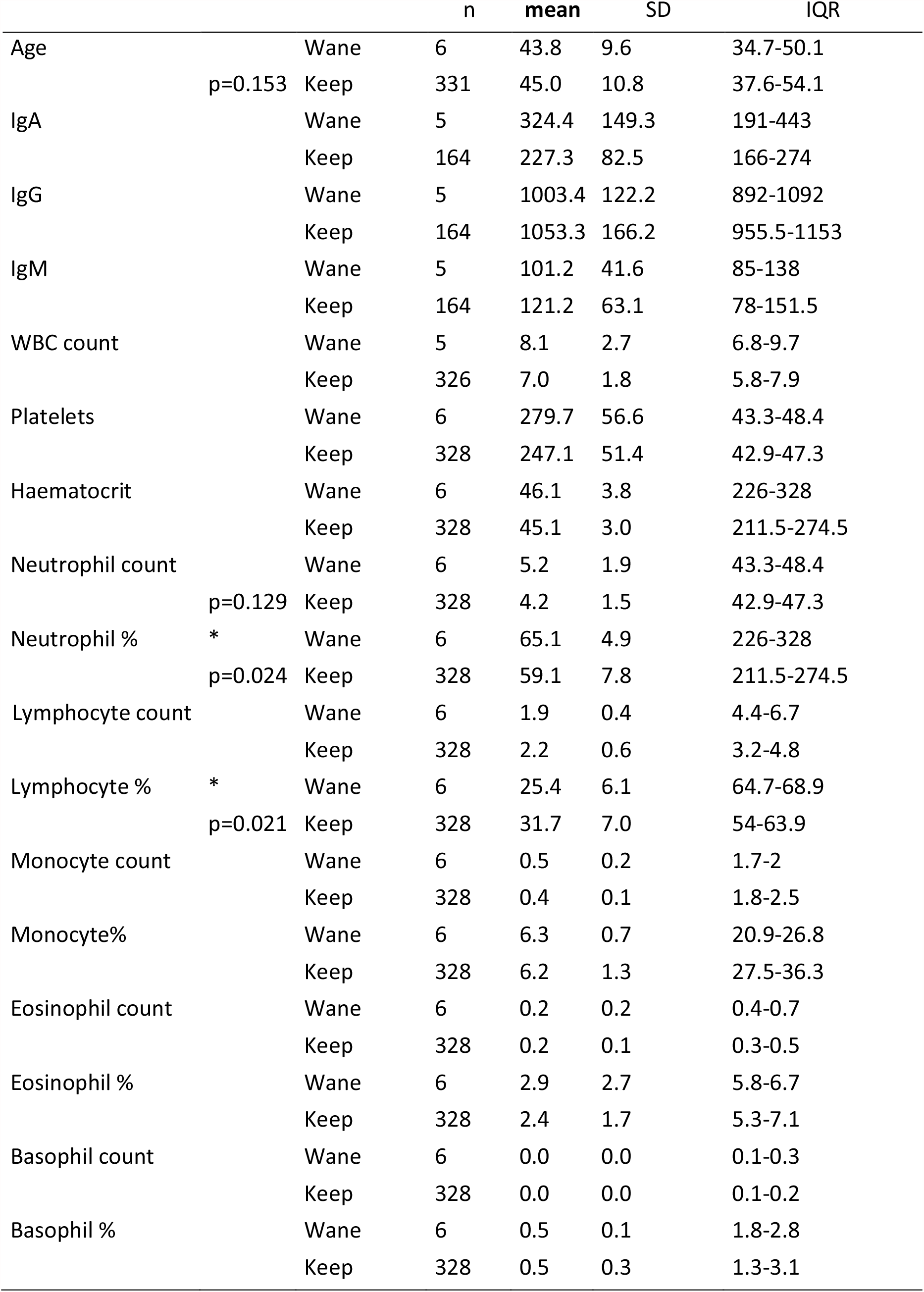
Age and laboratory data in donations keeping or waning antibodies Wane: present donation positive, next one, negative; keep: both positive

|  |  |  | n | mean | SD | IQR |
| --- | --- | --- | --- | --- | --- | --- |
| Age |  | Wane | 6 | 43.8 | 9.6 | 34.7-50.1 |
|  | p=0.153 | Keep | 331 | 45.0 | 10.8 | 37.6-54.1 |
| IgA |  | Wane | 5 | 324.4 | 149.3 | 191-443 |
|  |  | Keep | 164 | 227.3 | 82.5 | 166-274 |
| IgG |  | Wane | 5 | 1003.4 | 122.2 | 892-1092 |
|  |  | Keep | 164 | 1053.3 | 166.2 | 955.5-1153 |
| IgM |  | Wane | 5 | 101.2 | 41.6 | 85-138 |
|  |  | Keep | 164 | 121.2 | 63.1 | 78-151.5 |
| WBC count |  | Wane | 5 | 8.1 | 2.7 | 6.8-9.7 |
|  |  | Keep | 326 | 7.0 | 1.8 | 5.8-7.9 |
| Platelets |  | Wane | 6 | 279.7 | 56.6 | 43.3-48.4 |
|  |  | Keep | 328 | 247.1 | 51.4 | 42.9-47.3 |
| Haematocrit |  | Wane | 6 | 46.1 | 3.8 | 226-328 |
|  |  | Keep | 328 | 45.1 | 3.0 | 211.5-274.5 |
| Neutrophil count |  | Wane | 6 | 5.2 | 1.9 | 43.3-48.4 |
|  | p=0.129 | Keep | 328 | 4.2 | 1.5 | 42.9-47.3 |
| Neutrophil % | * | Wane | 6 | 65.1 | 4.9 | 226-328 |
|  | p=0.024 | Keep | 328 | 59.1 | 7.8 | 211.5-274.5 |
| Lymphocyte count |  | Wane | 6 | 1.9 | 0.4 | 4.4-6.7 |
|  |  | Keep | 328 | 2.2 | 0.6 | 3.2-4.8 |
| Lymphocyte % | * | Wane | 6 | 25.4 | 6.1 | 64.7-68.9 |
|  | p=0.021 | Keep | 328 | 31.7 | 7.0 | 54-63.9 |
| Monocyte count |  | Wane | 6 | 0.5 | 0.2 | 1.7-2 |
|  |  | Keep | 328 | 0.4 | 0.1 | 1.8-2.5 |
| Monocyte% |  | Wane | 6 | 6.3 | 0.7 | 20.9-26.8 |
|  |  | Keep | 328 | 6.2 | 1.3 | 27.5-36.3 |
| Eosinophil count |  | Wane | 6 | 0.2 | 0.2 | 0.4-0.7 |
|  |  | Keep | 328 | 0.2 | 0.1 | 0.3-0.5 |
| Eosinophil % |  | Wane | 6 | 2.9 | 2.7 | 5.8-6.7 |
|  |  | Keep | 328 | 2.4 | 1.7 | 5.3-7.1 |
| Basophil count |  | Wane | 6 | 0.0 | 0.0 | 0.1-0.3 |
|  |  | Keep | 328 | 0.0 | 0.0 | 0.1-0.2 |
| Basophil % |  | Wane | 6 | 0.5 | 0.1 | 1.8-2.8 |
|  |  | Keep | 328 | 0.5 | 0.3 | 1.3-3.1 |

ODs decayed as considering consecutive pairs of donations in 140 donors (56.0%, mean lapse between analysis, 9.5 weeks), rose in 54 (21.6%, mean lapse 11 weeks) cases and stayed +/-10% in 56 (22.4%, mean lapse 5.43 weeks) donors (Figure 1, Table 4). Donors whose ODs decayed were significantly younger (43 vs 47; p=0.042). The ones whose OD due to anti SARS-CoV-2 total antibodies stayed +/-10% had lower IgA as a trend (191.5mg/dL vs. 224.3 or 227.5 mg/dL; p=0.048) and a higher percent of basophils (0.6% vs. 0.5%; p=0.146)

**Table 4.**
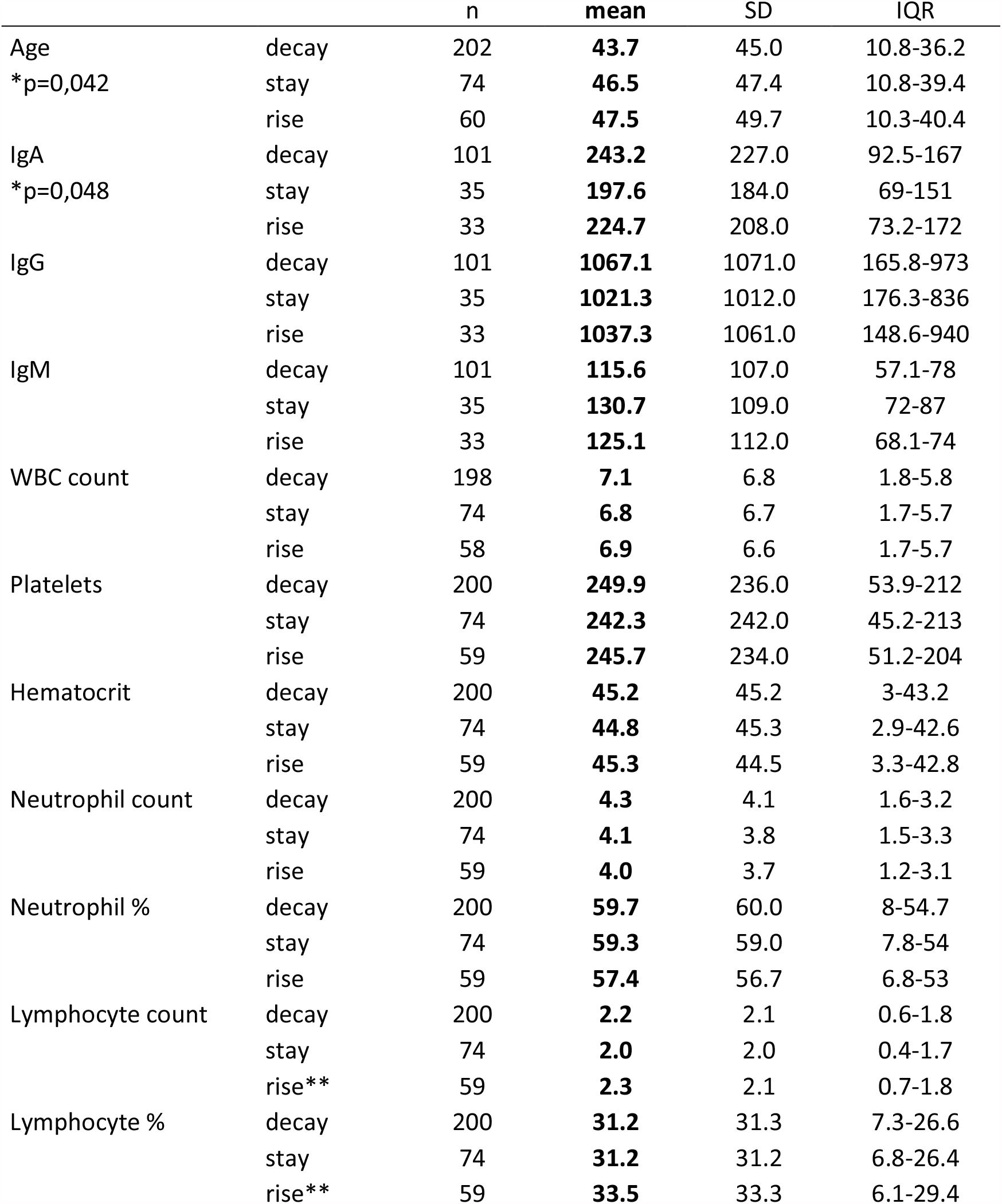
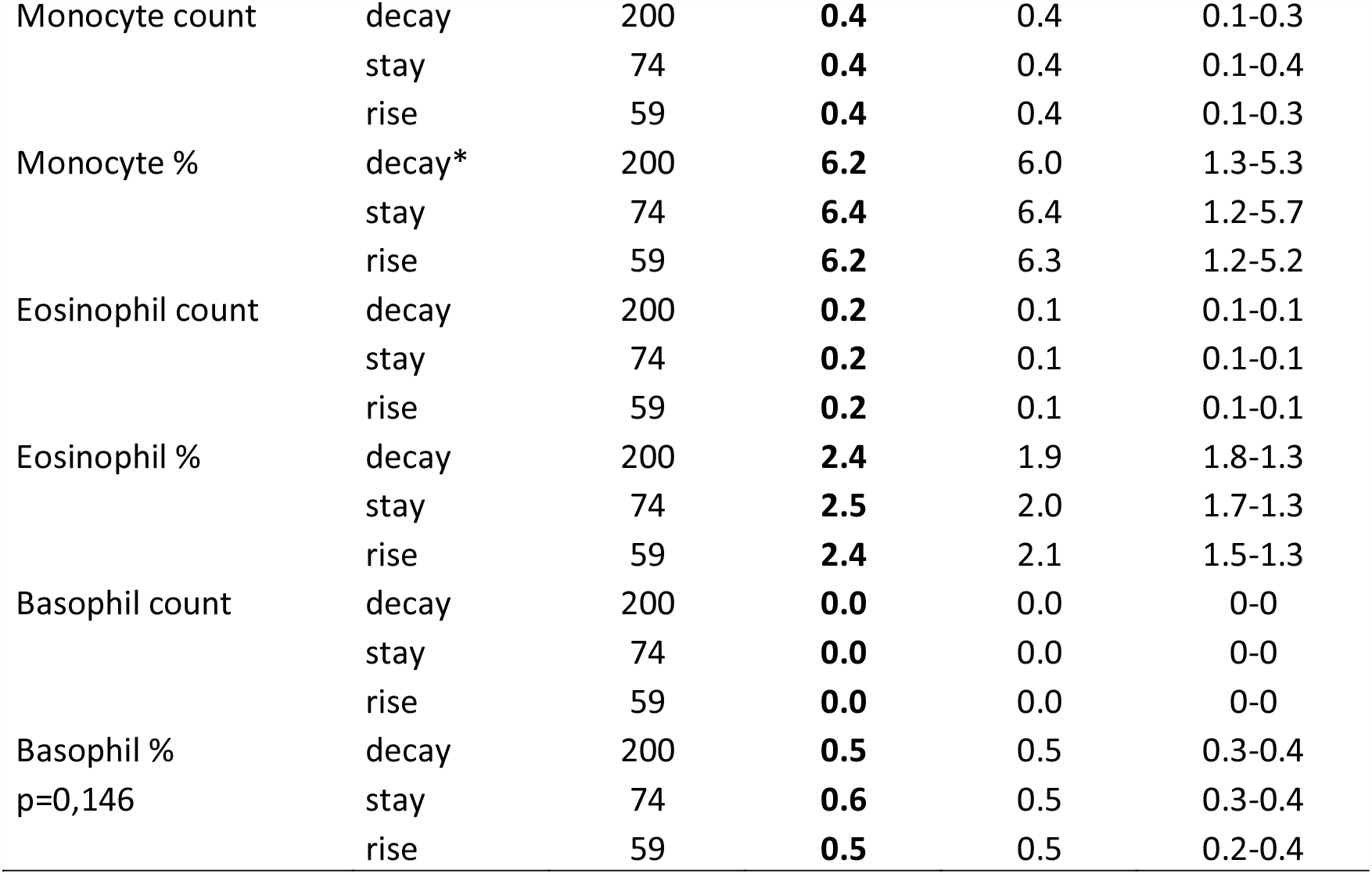
Age and laboratory donation data layered by OD observed changes in total anti SARS-CoV-2 antibodies (rise: the second determination was over 110% of the first; stay: OD of the second donation was 90%-110% of the first one; decay: OD of the second donation was under 90% of the first one)

When an age-stratified analysis was performed, some facts become noticeable as regarding just the ≥45 and <60 group: higher leukocytes arose as a significant difference in those who would wane (9.65 vs. 6.59; p=0.047), The difference in platelets is more evident as regarding just this group (327 vs. 231 p=0.019), the same happens with neutrophils both in absolute numbers (5.59 vs. 3.78; p=0.019) and percent (66.95 vs 58.25; p=0.014), lymphocyte count (1.86 vs 2.02; p=0.005), monocyte number (0.58 vs 0.41; p=0.028). Eosinophils were lower under 45: in under 30 group (0.02 vs 0.12 p= 0.049) and in ≥45 and <60 group (0.23 vs.0.15; p=0.034) (Supplementary Table 1).

## Discussion

There is little literature on SARS-CoV-2 antibody waning. Antibodies recognizing N protein of SARS-CoV-2 have been elsewhere reported to wane in the post infection phase while those against the S protein would persist over time [13]. Additionally, the antibody response would be robust in patients with severe infection and weaker in those with mild infection or asymptomatic individuals. Although 71% of repeat donors in this study were convalescent plasma donors, one of our limitations is that we cannot recall how many of them developed severe symptoms. Some reports at the beginning of pandemics pointed out that up to 40% asymptomatic cases could lose their antibodies after 6 weeks from acute infection [13] but our data don’t support that statement. More recent studies [1] demonstrate that durable serum antibodies would be granted by long-lived plasma cells. Our study confirms that most individuals would keep their antibodies for 10 months or longer. Our results agree as well to others reporting a high percentage of individuals (above 75%) keeping antibodies positive for at least 9 months even when a titer decline is patent [14]. That would mean at least 50 weeks after contagion so far they’re not allowed to donate until 1 month after resolution of symptoms.

Specific immunoglobulins would became detectable within 5-7 days post infection and seroconversion would happen after 10-14 days. Detectable levels of neutralizing antibodies against SARS-CoV-2 have been shown to start declining within three months of infection, especially among mild and asymptomatic cases [15]. Accordingly, one half (56.0%, Figure 1) of our donors exhibit a decay in their OD from total immunoglobulins anti SARS-COV-2 assay although most of them remain positive.

Antibody waning would be independent of blood group in our cohort. Specific anti SARS-CoV-2 immunoglobulin loss would be a rare event slightly more frequent in women and a decay would be more frequent in younger donors. Mature age would determine longer antibody half-life [14]. Ageing is one of the most important determinants in COVID-19 severity [16, 17] and it would have a role as well in humoral response keeping but larger cohorts and replication analyses should be performed to determine whether a an age-titer correlation exists because of the small rate of waning cases.

Lower lymphocyte and higher neutrophil percent might indicate future antibody waning. High neutrophil/lymphocyte ratios are somehow related to impaired immune responses with a high inflammatory component and cytokine storm [18]. Our findings would reinforce the hypothesis of a not so robust response that would weaken in the medium term. Some of these changes are more evident in mid-aged cases, an ageing effect cannot be discarded but the low number of waning cases, and the fact that none of them was older than 51 keeps us from further ascertainments, even when it points out a very interesting topic for future analyses.

Total, non-specific, high IgA together with higher platelet count and lower neutrophil count might precede antibody waning as well. It is well known that several infectious agents such as malaria have prothrombotic effects, which happen as well in bacterial sepsis [19]. It has been elsewhere reported an elevation of platelets and their activation and a lower number and activity of neutrophils in COVID-19 cases irrespective of their severity [20] accordingly to our data.

Secreted IgA is known to play a key role in SARS-CoV-2 immunity but very little is known on circulating IgA [21]. IgG in combination with IgM and IgA would have greater neutralization capacity than IgG alone, a broader repertoire of antibodies correlates with a stronger SARS-CoV-2 neutralization [22]. Initial IgA plasmablast’s response declines quickly, whereas IgA-producing plasma cells persist for years in mucosae [23]. It might explain that individuals mounting a strong IgA response would seem to loose antibodies, but it will be interesting to check out whether salivary IgA remains positive or not so far IgA antibodies are the major component of the neutralizing antibodies developed in response to SARS-CoV-2 infection [24]

Lower lymphocyte counts and higher neutrophils and as well higher seric IgA would help predict future negativization of antibodies. The vast majority of donors keep their total immunoglobulins anti SARS-CoV-2 positive for longer than 10 months. Ageing might have a protective effect against antibody waning but, given the small number of cases that become negative, more studies, or larger cohorts would be needed to confirm these facts.

## Data Availability

Fully anonimyzed raw data would be available after solicitude evaluation and approval by our Ethics Commitee

## Acknowledgements

LB and MCM conceived and designed the study. NO and AP acquired data, IP, AJ and MIG performed analysis and interpretation of laboratory data, MCM and LB drafted the article and revised it critically for important intellectual content, all authors provided final approval of the version to be submitted.

The authors thank to all blood donors for making this work possible by allowing research use of their samples by the Biobanco del Centro de Hemoterapia y Hemodonación de Castilla y León. They also thank the staff in charge of blood donation, and lab technicians for their efforts and from Roche Diagnostics International Ltd. for its support.

---

**Supplementary Table 1.**
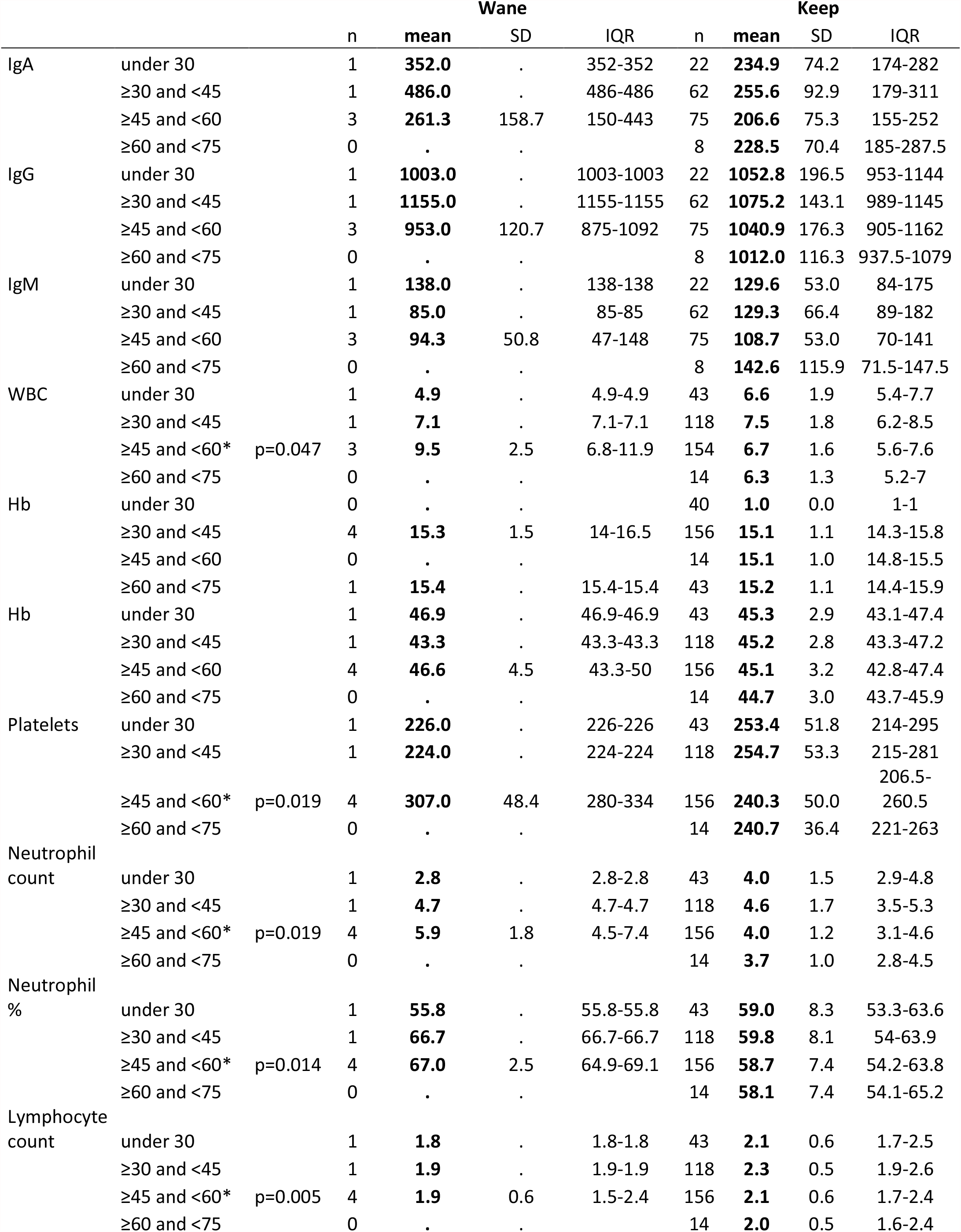
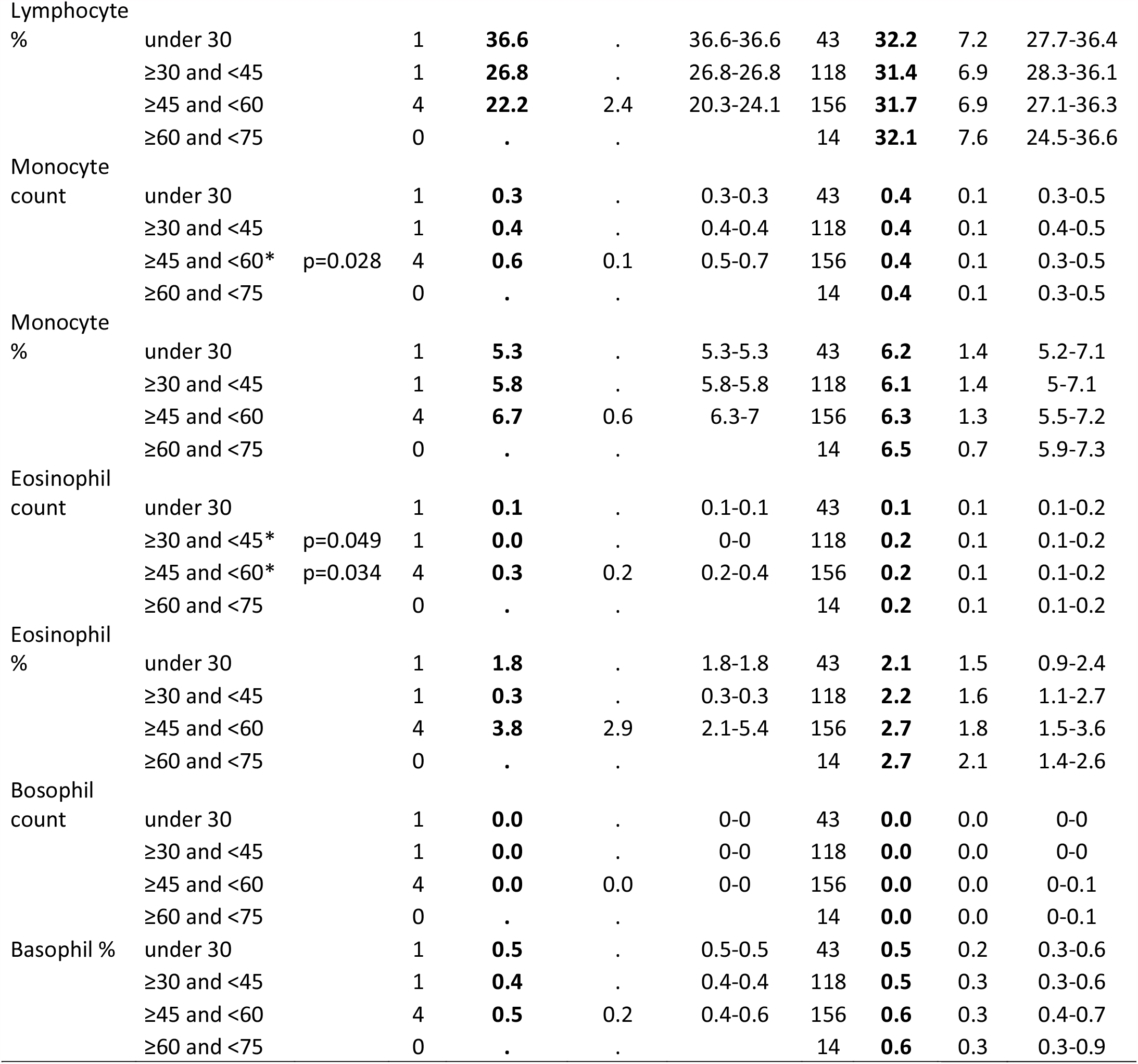
Laboratory data stratified by age in donations keeping or waning antibodies Wane: present donation positive, next one, negative; keep: both positive

